# Sex differences in Cerebral Small Vessel Disease: a systematic review and meta-analysis

**DOI:** 10.1101/2021.03.04.21252853

**Authors:** Lorena Jiménez-Sánchez, Olivia K. L. Hamilton, Una Clancy, Ellen V. Backhouse, Catriona R. Stewart, Michael S. Stringer, Fergus N. Doubal, Joanna M. Wardlaw

## Abstract

**Background:** Cerebral small vessel disease (SVD) is an important cause of acute ischemic stroke and vascular dementia. Several studies recruiting more males than females have reported sex differences regarding SVD incidence and severity, but it is unclear whether this reflects underlying sex-specific mechanisms or recruitment bias. This work aimed to systematically review and meta-analyze potential sex differences in SVD by assessing the male-to-female ratio (M:F) of recruited participants and incidence of SVD, risk factor presence, distribution and severity of SVD features.

**Methods:** Full text of 228 studies from four databases of recent systematic reviews on SVD and an independent search of MEDLINE were evaluated against inclusion and exclusion criteria (registered protocol: CRD42020193995). Data from participants with clinical or non-clinical presentations of SVD with radiological evidence of SVD were extracted. Sex ratios of total participants or SVD groups were calculated and differences in sex ratios across time, countries, SVD severity and risk factors for SVD were explored.

**Results:** Amongst 123 relevant studies (n = 36,910 participants) including 53 community-based, 67 hospital-based and 3 mixed studies, more males were recruited in hospital-based than in community-based studies (M:F = 1.16 (0.70) vs M:F = 0.79 (0.35), respectively; p <0.001). More males had moderate to severe SVD (M:F = 1.08 (0.81) vs M:F = 0.82 (0.47) in healthy to mild SVD; p <0.001), especially in stroke presentations where M:F was 1.67 (0.53). M:F of recent research (2015-2020) did not differ from that published pre-2015 and no geographical trends were apparent. There were insufficient sex-stratified data to explore M:F and risk factors for SVD.

**Conclusions:** Our results highlight differences in male-to-female ratios in SVD that may reflect sex-specific variability in risk factor exposures, study participation, clinical recognition, genuine SVD severity, or clinical presentation and have important clinical and translational implications.

## Introduction

Cerebral small-vessel disease (SVD) is a disorder of the brain small penetrating blood vessels leading to white and deep gray matter damage^1,2^, and is a major cause of stroke^3^ and/or dementia.^1^ Sex differences are well known in many vascular diseases^4^ but remain underexamined in SVD. Most SVD cases are sporadic, although there are rare monogenic types like cerebral autosomal dominant arteriopathy with subcortical infarcts and leukoencephalopathy (CADASIL), which is not sex-linked but males seem to be more severely affected than females.^5^ Several studies of sporadic SVD presenting as stroke have recruited more males than females and reported a higher age-adjusted incidence in males, but higher severity in females.^6^ On average, females are older at stroke onset, more likely to live alone and have more severe baseline deficits^7^, which could explain their increased pre-hospital delay, severity in first-ever acute stroke^8^ and post-stroke disability.^9^ These factors can affect females’ eligibility for stroke research studies, with a bias towards recruitment of milder strokes, and stroke treatment, as females are less likely to be treated with IV thrombolysis than males.^10^ Interestingly, females were more likely to refuse participation in stroke clinical trials than males independently of their age.^11^

We aimed to explore sex differences in SVD by assessing the sex ratio of participants with clinical or radiological evidence of SVD recruited to research studies. We assessed the incidence of SVD, the presence and distribution of risk factors for SVD and the severity of SVD features in males versus females.

## Methods

This work was performed in accordance with PRISMA guidelines. The protocol was registered on the PROSPERO database on July 2, 2020 (CRD42020193995).^12^

### Current databases

Articles from four recent systematic reviews^13–16^ that met our inclusion criteria (see below) were included. These systematic reviews provided a large publication sample that had already been screened against objective criteria, quality assessed and conducted according to PRISMA standards (Table 1).

**Table 1.** Systematic reviews.

| **Study<sup>15-18</sup>**<br>**(Primary author, year)** | **Title** | **Identified studies** | **Included studies** | **Total number of included participants in each review** |
| --- | --- | --- | --- | --- |
| Backhouse, 2017 | Early life risk factors for cerebrovascular disease. | 19,180 | 29 | 23,356 |
| Clancy, 2020 | Neuropsychiatric symptoms associate with cerebral small vessel disease: systematic review and meta-analysis. | 7,119 | 81 | 21,730 |
| Hamilton, 2020 | Cognitive impairment in sporadic cerebral small vessel disease: a systematic review and meta-analysis. | 8,562 | 69 | 6,908 |
| Stewart, 2020 | Associations between white matter hyperintensity burden, cerebral blood flow and transit time in small vessel disease: an updated meta-analysis. | 783 | 30 | 3,396 |

Identified studies refer to those found by search after duplicates were removed. Included studies refer to those examined for data extraction. The number of included studies of each systematic review do not include duplicated studies or populations.

### Search methods

To explore the most recent research including participants with SVD, an independent database search was also carried out. The search strategy was modified from a published protocol^16^ to identify studies including participants with clinical (stroke or cognitive presentations) or non-clinical presentations of sporadic or monogenic SVD (e.g. CADASIL). Stroke presentations included lacunar or subcortical stroke. Cognitive presentations included vascular cognitive impairment, either vascular mild cognitive impairment – VaMCI – or vascular dementia – VaD. Non-clinical presentations included radiological evidence of SVD – e.g. white matter hyperintensities (WMH), lacunes of presumed vascular origin, small subcortical infarcts or cerebral microbleeds (CMBs) on brain magnetic resonance imaging (MRI)^2^ – in the absence of clinical diagnosis (generally in community-dwelling populations), i.e. incidental ‘silent’ SVD. We searched MEDLINE through OVID for human studies published in English or Spanish from January, 1, 2015 to May, 26, 2020. The search strategy was as follows: Cerebral Small Vessel Diseases/ OR (small vessel disease or small vessel-disease or CSVD or SVD).ti.ab. OR stroke,Lacunar/ OR ((lesion* or hyperinten*) adj3 white matter).ti.ab. OR Leukoaraiosis/ OR lacune*.ti.ab. OR ((lacun* or subcort* or ischemi* or ischaemi* or silent or microscopic) adj3 lesion*).ti,ab.). Since the independent database search was designed as a sample to supplement all the studies collected from the 4 systematic reviews, only the most recent 150 journal articles among the 4,871 filtered results were examined. The electronic search was carried out on May, 26, 2020.

### Inclusion and exclusion criteria

Cross-sectional and longitudinal studies published in English or Spanish that considered clinical diagnosis of SVD or radiological markers for SVD were included. Review papers other than the included systematic reviews, editorials, communications, case reports, case series and conference abstracts were excluded. Studies about other neurodegenerative conditions (e.g. Parkinson’s disease, Alzheimer’s Disease, non-vascular or mixed dementia), inflammatory disorders (e.g. encephalitis/meningitis/vasculitis), single-sex populations (e.g.: pregnancy studies), genetic-based studies that only recruited from families, those that did not report proportions of males and females and those of acute ischemic stroke which did not stratify per stroke type (cortical or lacunar stroke) were excluded. To avoid possible confounding factors related to large vessel disease, studies that recruited participants based on cardiovascular events (e.g. heart failure) and diffuse cardiovascular disease (e.g. atherosclerosis) were excluded.

Where more than one study presented data on the same population, the study considering the most information about SVD clinical diagnosis, radiological markers or risk factors for SVD was selected.

### Data extraction

Screening, full-text review, and data extraction were independently carried out by five authors (L.J.S., O.K.L.H., E.V.B., U.C. and C.R.S.). Extracted data included the primary author, date of publication, country of recruited participants, study type (cross-sectional or longitudinal), clinical or non-clinical presentation of participants (including lacunar or subcortical stroke, subjective memory or cognitive complaints, VaMCI, VaD, presence and severity of WMH, lacunes, small subcortical infarcts, CMBs, silent brain infarcts, ICH or healthy participants), number of subjects, total sex ratio of participants, mean age of the population and sex-stratified mean age (both reported or calculated if data were available), stratified sex ratio by clinical diagnosis of SVD, radiological features of SVD or SVD score if provided. Since they are modifiable risk factors known to worsen SVD^17^, hypertension and current or ever-smoking data were recorded if available. Sex-stratified percentages of hypertension and smoking were calculated. Only baseline data were extracted in longitudinal studies.

### Statistical analysis

All analyses and plots were generated using R (version 3.2.3^18^). Sex ratios of total study participants or SVD groups of all the included studies were calculated and sex ratio per population type was compared. The principal summary measure was differences in the mean sex ratio.

Since recruitment can be affected by different factors across different settings, studies were classified into community-based, hospital-based, or mixed studies in which participants were recruited from both the community, and hospital and associated institutions. To investigate whether differences in sex ratios of the studies were influenced by the size of the recruited sample, a new variable was calculated: Δ sex ratio = |a constant of the global population sex ratio^19^ – sex ratio of each study|. Sizes of recruited populations were log-transformed due to their skewed distribution. The correlation of Δ sex ratio and the log-transformed size of the recruited populations per study type was then explored.

To explore trends across time and countries, studies were classified per year of publication and per country of recruited participants, respectively. To explore trends across severity and presentations of SVD, participants of the included studies were then classified into healthy to mild SVD and moderate to severe SVD (stroke presentations, cognitive presentations, moderate to severe non-clinical presentation and genetic SVD; detailed in Table 2).

**Table 2.** Study classification by SVD severity and presentation.

| **Group** | **Description** |
| --- | --- |
| Healthy to mild SVD | According to the definitions used in the original articles from which data was extracted: those defined as neurologically, functionally or cognitively healthy, community-dwelling individuals or participants with mild non-clinical presentation of SVD (with mild radiological features of SVD: deep or periventricular WMH, white matter lesions, vascular white matter disease, lacunes, leukoaraiosis, CMBs, silent brain infarcts or ICH). |
| Moderate to severe SVD | According to the definitions used in the original articles from which data was extracted: those with moderate or severe clinical or non-clinical presentations of SVD. This group includes stroke presentations, cognitive presentations, moderate to severe non-clinical presentations and genetic SVD. |
| Stroke presentations | Those first presenting with a lacunar or subcortical stroke or lacunar syndrome. Since cerebrovascular events can precede cognitive impairment, participants with both stroke and cognitive presentations of SVD (e.g. participants with lacunar stroke who also presented with VaD) were considered part of the stroke presentations group rather than the cognitive presentations group. |
| Cognitive presentations | Those presenting with self-reported and/or diagnosed cognitive impairment (subjective cognitive/memory complaints, subjective cognitive decline, VaMCI, VaD, subcortical ischemic vascular dementia or multi-infarct dementia). |
| Moderate to severe non-clinical presentations | Those presenting with incidental radiological features of SVD (deep or periventricular WMH, vascular white matter disease, lacunes, leukoaraiosis, CMBs, silent brain infarcts or ICH). |
| Genetic SVD | CADASIL |

Abbreviations: CADASIL = cerebral autosomal dominant arteriopathy with subcortical infarcts and leukoencephalopathy, CMB = cerebral microbleeds, ICH = intracerebral hemorrhage, SVD = cerebral small vessel disease, VaD = vascular dementia, VaMCI = vascular mild cognitive impairment, WMH = white matter hyperintensities.

For quantitative analyses, Shapiro-Wilk tests were used to check for data normality. Sex ratio and sex-stratified data were not normally distributed so non-parametric statistical tests were used. The Mann-Whitney-Wilcoxon test was used to explore comparisons between two groups and the Kruskal-Wallis test was used to explore comparisons between more than two groups. If the result of the Kruskal-Wallis test was significant, data were further analysed by pairwise Mann-Whitney-Wilcoxon followed by Bonferroni post-hoc correction. Correlation analyses were explored by calculating Spearman’s rank correlation coefficient. In text, data are presented as median (interquartile range, IQR). The significance threshold was set at p < 0.05.

### Quality assessment and publication bias

Quality assessment was carried out as in a previously published study^16^, rated on a scale from 0-8 according to STROBE guidelines. The median and IQR of the quality score of the included studies were calculated. To check sensitivity, meta-analyses were re-run excluding studies with quality scores lower than the median quality score of all included studies. Since very few studies have been published specifically on male-to-female ratios in SVD, publication bias was not assessed in this study.

### Data availability statement

Any data not published within the article can be shared by request from any qualified investigator.

## Results

Our work found 241 relevant journal articles from four systematic reviews and an independent search on MEDLINE through OVID. After filtering by language, full texts of 228 publications were assessed against exclusion criteria. Data were extracted and meta-analyzed from 123 studies that met the inclusion/exclusion criteria (n = 36,910 total participants, see supplementary references). Two studies explored genetic SVD (CADASIL) and 121 studies explored sporadic SVD. Study selection is detailed in figure 1 and the characteristics of the included studies are summarized in table S1. Studies were conducted from 1989 to 2020 in 23 countries across six continents (Europe = 43; Asia = 39; North America = 35; South America = 3; Australia = 2; Africa = 1).

**Figure 1.**
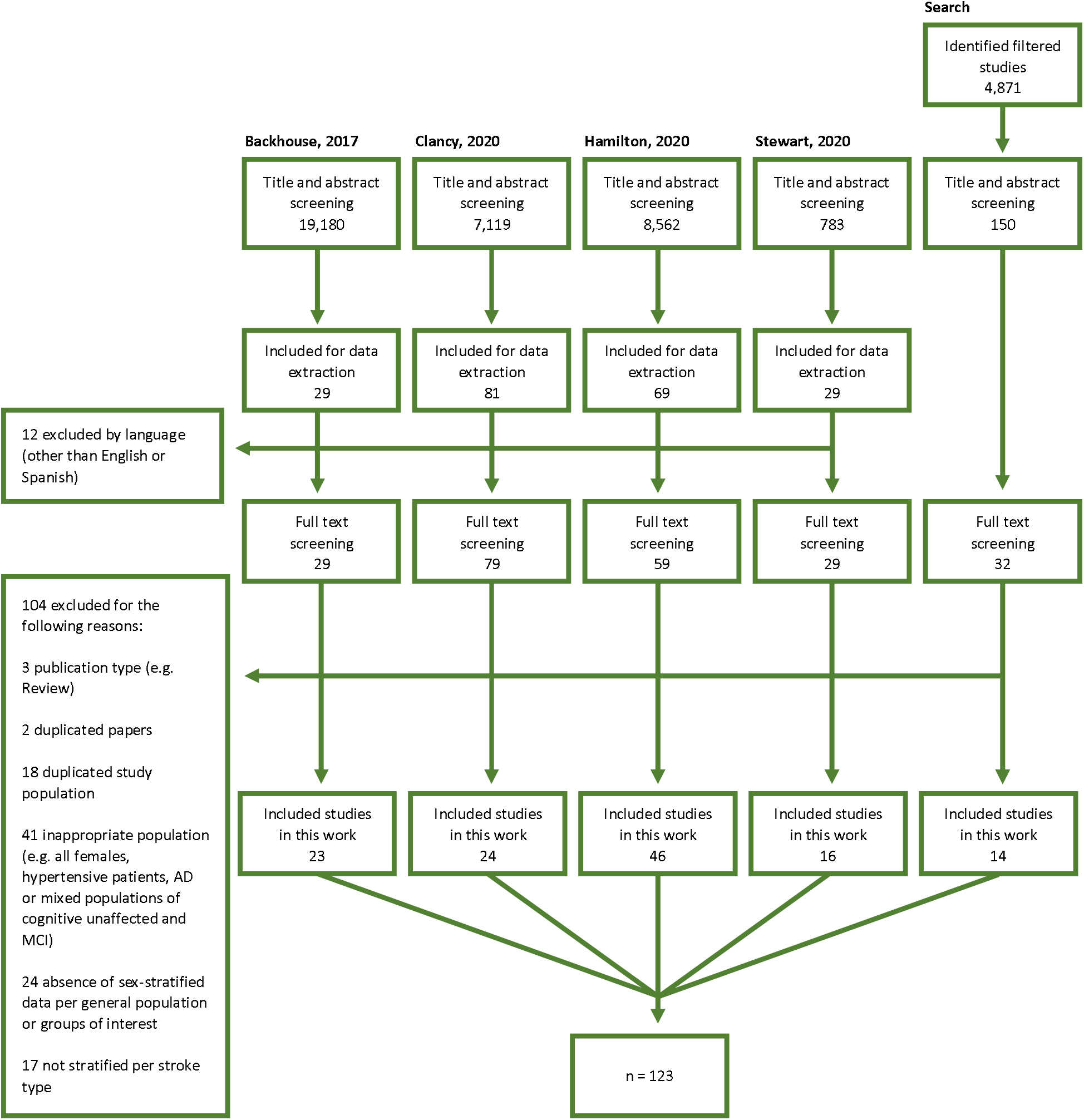
Study selection flow diagram. Abbreviations: AD = Alzheimer’s Disease, MCI = mild cognitive impairment.

None of the included studies reported both gender- and sex-stratified data or data regarding non-binary participants. Hence, for simplicity and without prejudice, sex ratios are referred to as male/female ratios.

### Trends across study settings

Our literature search retrieved 53 community-based (n = 29,323 total participants), 67 hospital-based (n = 7,337 total participants) and 3 mixed studies (n = 250 total participants). Global sex ratio of all included studies was 0.92 (0.65). Significant differences were found in sex ratios across study setting (H = 24.35, df = 2, p < 0.001). Sex ratio of hospital-based studies was greater when compared with community-based studies: 1.16 (0.70) vs 0.79 (0.35), respectively (p_corrected_ < .001; figure 2A). Considering that the mean age of the participants of the included studies was 67, sex ratio of community-based studies was closer to the expected general population sex ratio (0.89 in a 70-year old population^19^) than that of hospital-based studies.

**Figure 2.**
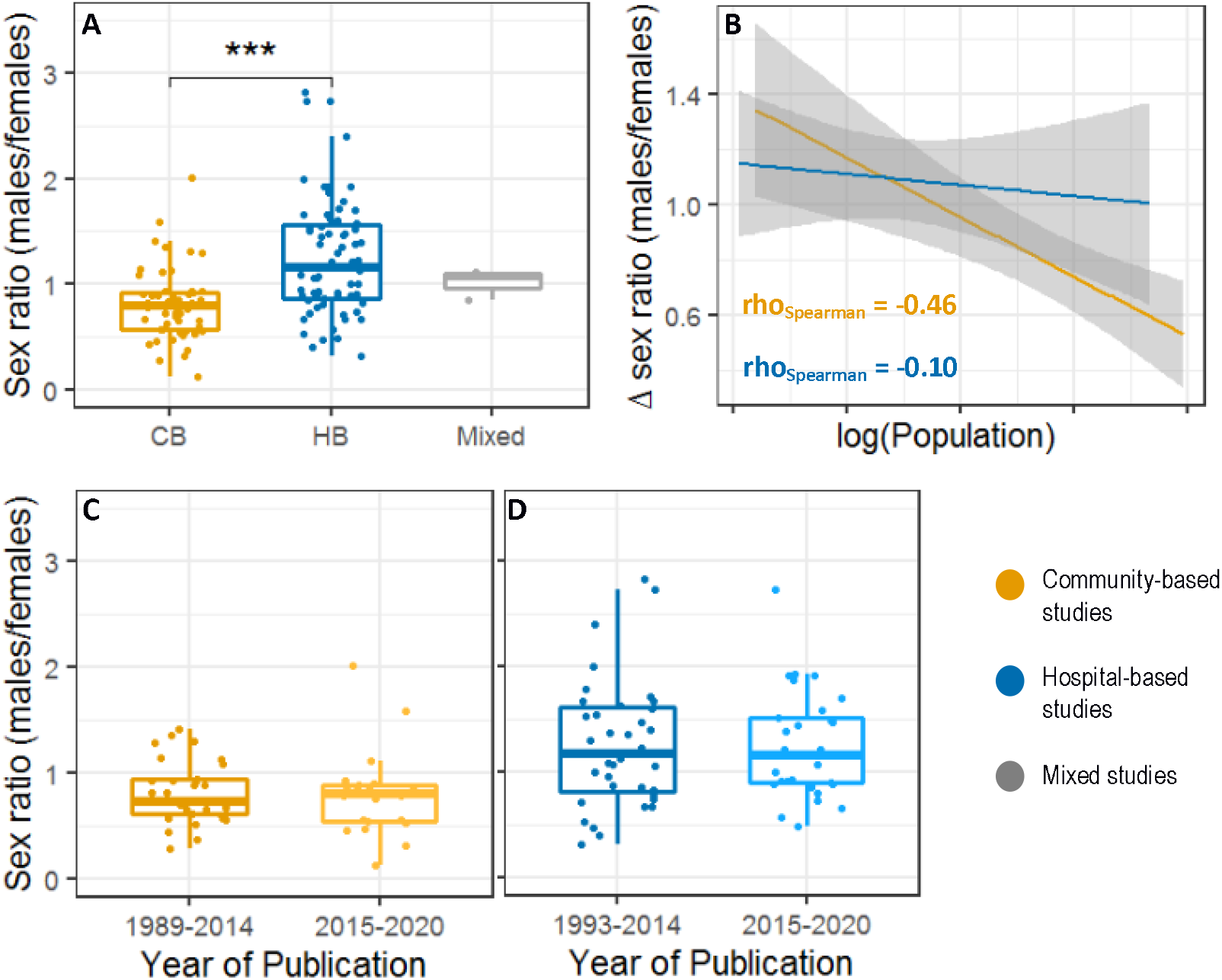
Sex ratio of SVD studies across study setting and time. (A) Comparison of sex ratios per study type. Significant differences were found between sex ratios of community-based (CB) and hospital-based (HB) studies (p_corrected_ < .001). (B) Correlation between the sex ratio difference and the size of the recruited sample. Δ sex ratio = |sex ratio of general population – sex ratio of each study|. Given that the mean age of the participants of the included studies was 67, general population sex ratio corresponds to 70-year old population (89 males per 100 females).^19^ There was a negative correlation between Δ sex ratio and the size of the population recruited in CB studies (yellow, p < 0.001) but not in HB studies (blue, p = 0.43). (C-D) Comparison of sex ratios across time. No significant differences were found between sex ratios of recent studies compared with those previously published considering all included studies (n_2015-2020_ = 53 vs n_1989-2014_ = 67, U = 1814, p = 0.75), (C) CB studies (n_2015-2020_ = 22 vs n_1989-2014_ = 31, U = 372, p = 0.58) or (D) HB studies (n_2015-2020_ = 31 vs n_1993-2014_ = 36, U = 551, p = 0.93). Abbreviations: CB = community-based, HB = hospital-based. *\*\*\* p < 0*.*001*

The effect of study size on sex ratio was different within community-based and hospital-based studies. The sex ratio of the studies was closer to the sex ratio of the general population when the sample size was greater in community-based studies (rho = −0.46, p < 0.001; figure 2B), but no effect was found in hospital-based studies (rho = −0.10; p = 0.43; figure 2B).

### Trends across time

Studies were classified per year of publication into recent (from 2015 to 2020) and previously published (until and including 2014) studies. Considering all the included studies, no significant differences were found between sex ratios of recent studies compared with those previously published (U = 1814, p = 0.75). This finding was consistent after classifying by study type (U = 372, p = 0.58 in community-based studies; U = 551, p = 0.93 in hospital-based studies; figure 3C,D). Mixed studies^137-139^ were not included in this analysis since only three were retrieved by our literature search, all published recently.

**Figure 3.**
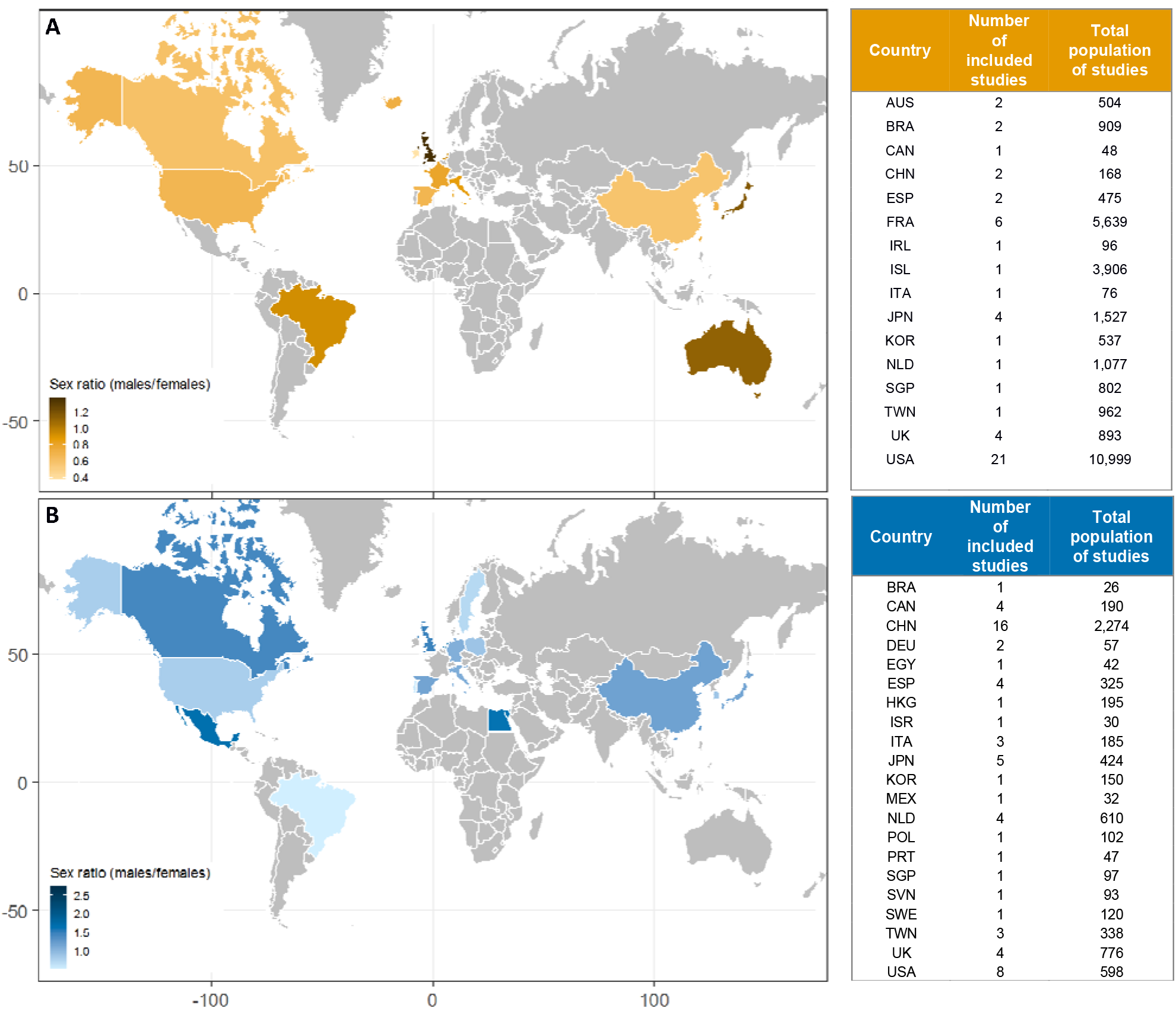
Sex ratio of SVD studies across the world. Colored world maps representing the mean sex ratio of the total number of participants of (A) community-based and (B) hospital-based studies. Darker shades in the color gradient correspond to higher sex ratios (i.e.: more males than females). The tables on the right specify the country of recruited participants, the number of included studies and the total population of included studies per study type. Neither multicentre nor mixed studies were represented in these maps. Abbreviations: AUS = Australia, BRA = Brazil, CAN = Canada, CHN = China, DEU = Germany, EGY = Egypt, ESP = Spain, FRA = France, HKG = Hong Kong, IRL = Ireland, ISR = Israel, ITA = Italy, JPN = Japan, KOR= Korea, MEX = Mexico, NLD = The Netherlands, POL = Poland, PRT = Portugal, SGP = Singapore, SVN = Slovenia, SWE = Sweden, TWN = Taiwan, UK = United Kingdom, USA = The United States of America.

### Trends across countries

Community-based and hospital-based studies were classified by country of recruited participants (figure 3). For clarity, studies that recruited participants from several countries^20–24^ and mixed studies were excluded.

Regarding community-based studies, the highest sex ratio was found in participants recruited from the United Kingdom (1.36 (0.19), four studies, n = 893) while participants recruited from the Republic of Ireland had the lowest sex ratio (0.37, one study, n = 96; figure 3A). The largest recruited population came from the United States of America (21 studies, n = 10,999 participants) with a median sex ratio of 0.67 (0.36). Regarding hospital-based studies, the highest sex ratio was found in participants recruited from Singapore (2.73, one study, n = 97) while participants recruited from Brazil had the lowest sex ratio (0.53, one study, n = 26; figure 3B). The largest recruited population came from China (16 studies, n = 2,274 participants) with a median sex ratio of 1.08 (0.48).

There were no obvious regional trends across countries for the sex ratio of the total number of participants for either community-based or hospital-based studies.

### Severity and presentation of SVD

The included studies enrolled a total of n = 25,972 healthy to mild SVD participants (no clinical presentation and mild radiological SVD features) and n = 10,938 moderate to severe SVD participants (clinical presentation and/or high radiological burden of SVD). Sex ratio in moderate to severe SVD was greater than in healthy to mild SVD: 1.08 (0.81) vs 0.82 (0.47; U = 3031.5, p < 0.001, figure 4A).

**Figure 4.**
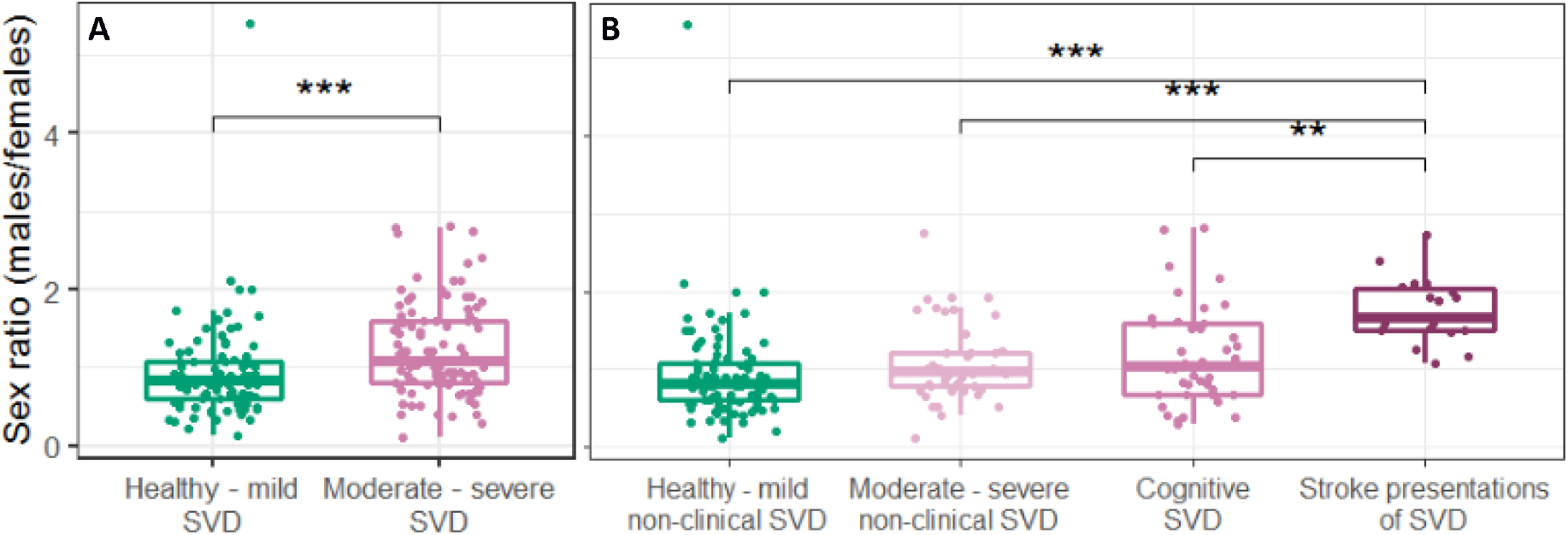
Sex ratio across SVD severity and presentation. Sex ratio of healthy to mild SVD compared with (A) moderate to severe SVD and (B) stratified moderate to severe SVD. Significant differences were found between SVD severity groups i.e. sex ratios of healthy to mild SVD and moderate to severe SVD (A; U = 3031.5, p < 0.001). Significant differences were also found between SVD presentation groups (H = 36.58, df = 3, p < 0.001) i.e. stroke presentations of SVD compared with healthy to mild non-clinical SVD, moderate to severe non-clinical SVD or cognitive SVD (B; p_corrected_ < .001, p_corrected_ < .001, p_corrected_ = .003, respectively). *\*\* p < 0*.*01, *** p < 0*.*001*

Moderate to severe SVD participants were further classified into cognitive or stroke presentations or non-clinical SVD (figure 4B). Insufficient data were available for genetic SVD (CADASIL^22,25^), so these two studies were excluded from this analysis. Significant differences were found in sex ratios across SVD presentation (H = 36.58, df = 3, p < 0.001). Participants with stroke presentations showed the highest sex ratio, 1.67 (0.53), greater when compared with healthy to mild SVD (0.82 (0.47), p_corrected_ < .001), cognitive SVD (1.03 (0.91), p_corrected_ = .003), and moderate to severe non-clinical SVD (0.96 (0.44), p_corrected_ < .001).

Given that community-based studies recruited a vast majority of healthy participants and that they presented lower sex ratios, the same severity analysis was performed in hospital-based studies only. The trends reported in all included studies were consistent within this group: sex ratio across SVD severity groups (U = 1239.5, p < 0.001) - 1.26 (0.87) in moderate to severe SVD vs 0.90 (0.58) in healthy to mild non-clinical SVD; sex ratio across SVD presentation groups (H = 21.82, df = 3, p < 0.001) - 1.67 (0.55) in stroke presentations vs 0.90 (0.58) in healthy to mild non-clinical SVD (p_corrected_ < .001), 1.11 (0.81) in cognitive SVD (p_corrected_ = .037) and 1.13 (0.87) in moderate to severe non-clinical SVD (p_corrected_ = .02).

### Age and risk factors for SVD

Only 10 studies (2,953 total participants) provided sufficient data to calculate the sex-stratified age of participants. The median age in total recruited males was compared with that of total recruited females. There was no significant difference between the two groups: 63.78 (9.71) in males vs 64.45 (13.71) in females (U = 49.5, p > 0.99).

Only two studies^66,74^ allowed the calculation of sex-stratified data on risk factors for SVD (hypertension and ever smoking), so data were insufficient to perform further analyses.

### Quality assessment

The median study quality score was 5.5 (1). As a sensitivity analysis, quantitative analyses were re-run excluding all studies with a quality score < 5.5/8. All the trends observed in the total included studies were consistent in the subset of higher-quality studies (score ≥ 5.5/8; table S2).

## Discussion

This meta-analysis of 123 studies (n = 36,910 total participants, see supplementary references) evidences sex differences in SVD across study settings, SVD severity and presentation. A greater male-to-female ratio was found in hospital-based compared to community-based studies (figure 2). No differences were found between sex ratios of recent (2015-2020) and previously published studies (1989-2014), independently of the study setting (figure 2C,D). No regional trends were apparent in community-based or hospital-based studies (figure 3). The sex ratio was greater in moderate to severe SVD, particularly in stroke presentations when compared with healthy to mild SVD (figure 4).

The different sex ratios found between community-based and hospital-based studies may be due to differences in recruitment. Typically, females are older and have greater levels of disability at stroke onset^8^, which may affect study eligibility. For example, ischemic stroke patients older than 80 years have higher rates of disability following thrombolysis treatment^26^ and are less likely to be recruited into stroke trials.^27^ Therefore, our results may reflect recruitment bias towards younger and less disabled patients, likely males. Furthermore, women with stroke often present with non-traditional symptoms like altered mental status^28^, which could be overlooked or misdiagnosed in the clinical setting. ^29,30^ Sex differences in clinical presentations are also present in dementia^31^ but none of the included studies reported these in VaCI or VaD. Moreover, informal carers of dependent persons in the UK are more likely to be middle-aged women with multiple roles until later life (70+).^32^ Thus, females may be reluctant to participate in studies due to care responsibilities or may normalize their early symptoms while providing care. However, caregiving roles vary by country^33^, socioeconomic status and culture of care.^34^ This might explain why more females seemed to participate in Chinese hospital-based studies compared with the UK or Canada (figure 3B) since Chinese males are traditionally the predominant caregivers for older parents.^35^ Interestingly, some of the aforementioned factors that may alter female recruitment to SVD studies have recently been highlighted as contributors to lower enrolment of women in stroke clinical trials.^36^ It could also be that SVD is more prevalent and/or severe in males than in females, leading males to be more likely participants in studies investigating severe SVD. In support of this, male-sex was an independent predictor of severity of SVD in an adjusted analysis, albeit in a 62 % male population.^17^ Similarly, a greater prevalence of stroke, higher cognitive impairment and cerebral atrophy have been reported in men with CADASIL.^37^ Sex differences can be driven by sex-specific biological factors e.g. sexual dimorphism in endothelial function.^38^ In premenopausal females, oestrogens enhance endothelial production of vasodilator factors.^39^ This may explain young males having greater vasoconstrictor tone compared to pre-menopausal females^38^ and male endothelial function becoming suboptimal under certain insults. No differences in age between recruited males and females were found, although only 10 studies allowed the calculation of sex-stratified age of participants so their results may not be representative. Different lifestyle-related risk factors could also contribute to the sex-specific severity of SVD, e.g.: utilization of preventative health care services, smoking or hypertension. For example, fewer men are willing to participate in skin cancer screening.^40^ Additionally, the prevalence of smoking and hypertension is higher among males in most countries^41,42^, varying with race.^43^ These factors were more strongly associated with the risk of any stroke type in women compared with men in a recent study.^44^ Our work found insufficient data to analyse sex-specific risk factor effects driving the sex ratio difference in SVD severity and presentation.

The unequal sex ratios found here may be explained by factors with different contributions across different settings, evidenced by the different effect of study size on sex ratio within community-based and hospital-based studies (figure 2B), or in the context of higher SVD severity and stroke presentations. The fact that no significant differences were found between sex ratios of recent and earlier studies (figure 2C,D) may indicate that the same factors have been playing a role throughout time.

The implications for future research and clinical practice are varied and important. This work shows there is a lack of sex-stratified data, previously reported in brain structural studies^45^ and aging research^46^, that may hamper translational research and more personalized care across the lifespan. Thus, there is a need for reporting and analyzing results by sex, especially when biological factors, treatments or social disparities may differ between sexes.^47^ This matter has been addressed recently^48,49^ in support of the Sex and Gender Equity in Research (SAGER) guidelines^50^ and the European Commission second report on Gendered Innovations^51^, which provides guidance for researchers to incorporate sex, gender and intersectional analysis across several research topics. Future studies should also identify and avoid recruitment bias, explore whether SVD is more frequently underestimated or misdiagnosed in females and investigate possible reasons why males might be more severely affected. Larger sample sizes may help to reduce sampling variability at least within community-based studies with a majority of functionally healthy individuals (figure 2B). If the disease in females is going unrecognized, doctors and the public could be educated to better recognize atypical symptoms in females. If males are more severely affected or exposed to certain lifestyle factors, trials may need to target drivers of males’ vulnerability and health promotion campaigns could be designed to have more impact on males.

This study had several limitations. First, the pooled mean age for total participants per population/severity/presentation subtype was not extracted or calculated, which would have helped to understand the epidemiology of each group. Second, this study did not examine the functional status of participants with SVD, which may be heavily impacted by eligibility criteria, and result in the exclusion of females who are more functionally disabled. Third, some risk factors and their differences between sexes were not explored (e.g.: lower educational attainment, associated with increased risk of SVD on neuroimaging in later life).^13^ Fourth, this review relied on individual studies’ criteria for SVD severity. Future explorations could investigate the heterogeneity between study criteria and make further standardisation efforts. There were additional limitations of the included literature, such as the scarcity of sex-stratified data to explore age and risk factors for SVD. Additionally, the available data only allowed the investigation of sex and not gender differences, while it is possible that both may have different influences on health and disease. Finally, the studies retrieved by our search were mostly from industrialized countries (figure 3), so our results might not fully represent other populations.

To the best of our knowledge, this is the first systematic review and meta-analysis to explore sex differences in SVD. A broad approach was taken to capture changes across time, study settings, different cultural or ethnic groups, SVD severity and presentation. The included studies were conducted from 1989 to 2020, recruited 36,910 participants from the community and/or hospitals and associated institutions in 23 countries across six continents, and explored a wide range of SVD radiological features, signs, and symptoms. Our results highlight sex-specific variability in study participation, SVD severity and clinical presentation. These findings are relevant for future research and clinical practice, but more work is needed to unmask sex-specific biological and social disparities and to disentangle their contributions to sex differences in SVD. Further clarity could be sought through stroke and dementia registries, audit data and population-based epidemiological studies, which are all less prone to male/female recruitment bias.

## Data Availability

Any data not published within the article can be shared by request from any qualified investigator.

## Abbreviations

CADASIL: cerebral autosomal dominant arteriopathy with subcortical infarcts and leukoencephalopathy
CMBs: cerebral microbleeds
MRI: magnetic resonance imaging
ICH: intracerebral hemorrhage
SVD: cerebral small vessel disease
VaD: vascular dementia
VaCI: vascular cognitive impairment
WMH: white matter hyperintensities

## Author contributions

L.J.S. carried out the independent literature search, extracted the data, performed the meta-analyses and drafted the manuscript. O.K.L.H., E.V.B., U.C. and C.R.S. carried out the literature search of their corresponding systematic reviews and provided their databases, reviewed and edited the manuscript. M.S.S. co-supervised one of the systematic reviews (conducted by C.R.S.), checked and edited the manuscript. F.N.D. co-supervised one of the systematic reviews (conducted by U.C.), checked and edited the manuscript. J.M.W. conceived and managed the project, designed the protocol, checked the search strategy, supervised the contributing meta-analyses, reviewed uncertain articles, advised on the meta-analysis and interpretation of data, and reviewed and edited the manuscript. The final draft of the manuscript was approved by all authors.

## Declaration of interests

The authors declare no conflict of interest.

## Funding

L.J.S. is a Translational Neuroscience PhD student funded by Wellcome (108890/Z/15/Z). O.K.L.H. is a Translational Neuroscience PhD student funded by the College of Medicine and Veterinary Medicine at the University of Edinburgh. U.C. is funded by a Chief Scientist Office of Scotland Clinical Academic Fellowship (CAF/18/08) and Stroke Association Princess Margaret Research Development Fellowship (2018). E.V.B. is funded by the Sackler Foundation, the Stroke Association, British Heart Foundation and Alzheimer’s Society through the R4VaD Study. M.S.S. is funded by the Fondation Leducq (ref no. 16 CVD 05) and EU Horizon2020 (PHC-03-15, project No 666881, ‘SVDs@Target’) and the MRC UK Dementia Research Institute at the University of Edinburgh (UK DRI LTD, funded by the UK Medical Research Council, Alzheimer’s Society and Alzheimer’s Research UK). F.N.D. is funded by a Stroke Association Garfield Weston Foundation (TSALECT 2015/04) Senior Clinical Lectureship and NHS Research Scotland. J.M.W. is funded by the Stroke Association, British Hearth Foundation, Row Fogo Charitable Trust, Fondation Leducq (Perivascular Spaces Transatlantic Network of Excellence), and EU Horizon 2020 (SVDs@Target) and the MRC UK Dementia Research Institute at the University of Edinburgh. All authors hold grants from government/charitable agencies. The funding sources had no role in the study design, execution, analysis, interpretation of the data, decision to publish or preparation of the manuscript.

## Supplementary information

Please find supplementary tables (S1, S2) and references in the Supplementary Material file.

## Notes

### Competing Interest Statement

The authors have declared no competing interest.

### Clinical Protocols

https://www.crd.york.ac.uk/prospero/display_record.php?RecordID=193995&VersionID=1392786

### Author Declarations

This work reviews findings from previously published studies. Thus, no additional ethics committee approvals were required.

