## Supplementary Material for "Sex differences in Cerebral Small Vessel Disease: a systematic review and meta-analysis"

**Supplementary Materials**

**Supplementary Table S1. Characteristics of all included studies.**

**Supplementary Table S2. Results of quantitative tests excluding studies with quality scores < 5.5/8.**

**Supplementary references.**

**Supplementary Table S1. Characteristics of all included studies.**

| **Study (Primary author, year)** | **Community-based (C), hospital-based (H) or mixed (M)** | **Country of recruited participants** | **SVD features of selected participants (original definitions used)** | **Number of total participants** | **Mean age of total participants** | **Males** | | | | **Females** | | | |
| --- | --- | --- | --- | --- | --- | --- | --- | --- | --- | --- | --- | --- | --- |
|  |  |  |  |  |  | **n** | **Mean age** | **% Hypertension** | **% Ever smoking** | **n** | **Mean age** | **% Hypertension** | **% Ever smoking** |
| Kim, 2020^2^ | C | USA | Healthy participants  MCI | 38 | 68.31 | 7  11 | NA | NA | NA | 14  6 | NA | NA | NA |
| Cedres, 2019^3^ | C | Spain | Healthy participants | 416 | 58.5 | 190 | NA | NA | NA | 226 | NA | NA | NA |
| Dolui, 2019^4^ | C | USA | Healthy participants | 497 | 53.21 | 224 | NA | NA | NA | 273 | NA | NA | NA |
| Legdeur, 2019^5^ | C | USA | Healthy participants | 141 | 94.3 | 45 | NA | NA | NA | 96 | NA | NA | NA |
| Mishra, 2019^6^ | C | France | Minimal SVD  Extensive SVD | 580 | 64.75 | 98  109 | NA | NA | NA | 223  150 | NA | NA | NA |
| Puzo, 2019^7^ | C | USA | Healthy participants | 465 | 68.9 | 146 | NA | NA | NA | 319 | NA | NA | NA |
| Staffaroni, 2019^8^ | C | USA | Healthy participants | 161 | 69.9 | 73 | NA | NA | NA | 88 | NA | NA | NA |
| Tsapanou, 2019^9^ | C | USA | Healthy participants | 562 | 54 | 243 | NA | NA | NA | 319 | NA | NA | NA |
| Yao, 2019^10^ | C | Japan | Healthy participants | 259 | 68.4 | 122 | NA | NA | NA | 137 | NA | NA | NA |
| Croall, 2018^11^ | C | UK | Severe SVD (with confirmed LS) | 70 | 69.45 | 43 | NA | NA | NA | 27 | NA | NA | NA |
| Kuriyama, 2018^12^ | C | Japan | Controls  Deep WMLs Faz 1  Deep WMLs Faz 2  Deep WMLs Faz 3 | 280 | 70.8 | 45  92  40  10 | NA | NA | NA | 23  42  22  6 | NA | NA | NA |
| Puglisi, 2018^13^ | C | Italy | WMLs Faz 1  WMLs Faz 2  WMLs Faz 3 | 76 | 72.5 | 7  15  13 | NA | NA | NA | 13  17  11 | NA | NA | NA |
| Shokouhi, 2018^14^ | C | USA | Controls  MCI | 265 | 64.0 | 19  74 | NA | NA | NA | 61  111 | NA | NA | NA |
| Van Rooden, 2018^15^ | C | Netherlands and USA | Controls  Subjective cognitive decline | 67 | 68.0 | 17  7 | NA | NA | NA | 25  18 | NA | NA | NA |

| Bahrani, 2017^16^ | C | USA | Healthy participants | 26 | 77.8 | 3 | 77.0 | NA | NA | 23 | 77.8 | NA | NA |
| --- | --- | --- | --- | --- | --- | --- | --- | --- | --- | --- | --- | --- | --- |
| Squarzoni, 2017^17^ | C | Brazil | Controls  Silent vascular brain lesions | 234 | 73.91 | 88  25 | NA | NA | NA | 89  32 | NA | NA | NA |
| Shi, 2017^18^ | C | China | Healthy participants. | 69 | 70.78 | 24 | NA | NA | NA | 45 | NA | NA | NA |
| Xu, 2017^19^ | C | Singapore | Controls  1 CMB  Multiple CMB | 802 | 70.3 | 233  67  69 | NA | NA | NA | 289  95  49 | NA | NA | NA |
| Chung, 2016^20^ | C | Taiwan | Healthy participants | 962 | 62.5 | 425 | NA | NA | NA | 537 | NA | NA | NA |
| Promjunyakul, 2016^21^ | C | USA | Healthy participants | 82 | 84 | 20 | NA | NA | NA | 62 | NA | NA | NA |
| Vemuri, 2015^22^ | C | USA | Controls  Vascular pathology | 267 | 76.67 | 93  48 | NA | NA | NA | 85  41 | NA | NA | NA |
| Yamawaki, 2015^23^ | C | Japan | Mild DWMH Faz 0-1  Moderate DWMH Faz 2  Severe DWMH Faz 3 | 688 | 76.5 | 198  68  37 | NA | NA | NA | 240  108  37 | NA | NA | NA |
| Annweiler, 2014^24^ | C | France | Subjective memory complaint | 133 | 71.6 | 75 | NA | NA | NA | 58 | NA | NA | NA |
| Mortamais, 2014^25^ | C | France | Low WMLs < 0.3 ml  Mild WMLs 0.3-1.5 ml  Severe WMLs > 1.5 ml | 500 | 71 | 77  86  72 | NA | NA | NA | 95  85  85 | NA | NA | NA |
| Sarabia-Cobo, 2014^26^ | C | Spain | MCI | 59 | 77.80 | 20 | NA | NA | NA | 39 | NA | NA | NA |
| Sims, 2014^27^ | C | USA | Healthy participants | 172 | 64.43 | 99 | NA | NA | NA | 73 | NA | NA | NA |
| Sun, 2014^28^ | C | China | Controls  Mild WMLs | 99 | 64.11 | 16  22 | NA | NA | NA | 33  28 | NA | NA | NA |
| Wiegman, 2014^29^ | C | USA | Controls  CMBs | 243 | 84.50 | 51  23 | NA | NA | NA | 125  44 | NA | NA | NA |
| Farfel, 2013^30^ | C | Brazil | Healthy participants | 675 | 74.0 | 322 | NA | NA | NA | 353 | NA | NA | NA |

| Minn, 2013^31^ | C | South Korea | Controls  WM changes or SI | 537 | 63 | 122  99 | NA | NA | NA | 219  97 | NA | NA | NA |
| --- | --- | --- | --- | --- | --- | --- | --- | --- | --- | --- | --- | --- | --- |
| Nebes, 2013^32^ | C | USA | Controls  WMH | 66 | NA | 15  9 | NA | NA | NA | 25  17 | NA | NA | NA |
| Valdés-Hernández, 2013^33^ | C | UK | Healthy participants | 634 | 72.7 | 337 | NA | NA | NA | 297 | NA | NA | NA |
| Bartley, 2012^34^ | C | Ireland | Controls  Subjective memory complaints | 96 | 62.5 | 11  15 | NA | NA | NA | 33  37 | NA | NA | NA |
| Salarirad, 2011^35^ | C | UK | Healthy participants | 106 | 78.3 | 62 | NA | NA | NA | 44 | NA | NA | NA |
| Stewart, 2011^36^ | C | France | Healthy participants | 1,793 | 72.4 | 710 | NA | NA | NA | 1,083 | NA | NA | NA |
| Villeneuve, 2011^37^ | C | Canada | Controls  MCI with confluent WMLs | 48 | 72.05 | 7  11 | NA | NA | NA | 20  10 | NA | NA | NA |
| Qiu, 2010^38^ | C | Iceland | Controls  CMBs | 3,906 | 76 | 1,399  246 | NA | NA | NA | 2,057  204 | NA | NA | NA |
| Godin, 2009^39^ | C | France | Healthy participants | 1,792 | 72.4 | 708 | NA | NA | NA | 1,084 | NA | NA | NA |
| Anderson, 2008^40^ | C | Australia | Controls  First-ever lacunar syndrome | 60 | 68.63 | 16  16 | NA | NA | NA | 14  14 | NA | NA | NA |
| Miranda, 2008^41^ | C | Multicentre | LA without memory impairment  LA with memory impairment | 638 | 74.1 | 107  180 | NA | NA | NA | 132  219 | NA | NA | NA |
| Christensen, 2007^42^ | C | Australia | Healthy participants | 444 | 62.64 | 231 | 62.65 | NA | NA | 213 | 62.63 | NA | NA |
| Schretlen, 2007^43^ | C | USA | Healthy participants | 177 | 35.97 | 85 | 62.1 | NA | NA | 92 | 58.8 | NA | NA |
| Au, 2006^44^ | C | USA | Non-large WMH  Large WMH | 1,819 | 61.15 | 745  109 | NA | NA | NA | 834  131 | NA | NA | NA |
| Elkins, 2006^45^ | C | USA | Healthy participants | 3,622 | 75.06 | 1,524 | NA | NA | NA | 2,098 | NA | NA | NA |
| Wright, 2005^46^ | C | USA | Healthy participants | 259 | 64.8 | 116 | NA | NA | NA | 143 | NA | NA | NA |
| Deary, 2003^47^ | C | UK | Healthy participants | 83 | 78 | 47 | 78 | NA | NA | 36 | 78 | NA | NA |
| Dufouil, 2003^48^ | C | France | No lesion or mild WMH  Moderate WMH  Severe WMH | 841 | 69 | 145  149  58 | NA | NA | NA | 175  229  85 | NA | NA | NA |
| Tsukishima, 2001^49^ | C | Japan | Controls  With WML  With SI  Both WML and SI | 300 | 72.9 | 112  7  15  11 | NA | NA | NA | 121  8  20  6 | NA | NA | NA |
| De Groot, 2000^50^ | C | Netherlands | Controls  WMLs | 1,077 | 72.21 | 22  500 | NA | NA | NA | 32  523 | NA | NA | NA |
| Liao, 1997^51^ | C | USA | Controls  Mild WMLs  Moderate WMLs  Severe WMLs | 1,921 | 62 | 97  372  196  106 | Black males: 62  White males: 63 | Black males: 56  White males: 38 | Black males: 69  White males: 76 | 183  586  248  133 | Black females: 61  White females: 63 | Black females: 68  White females: 31 | Black females: 69  White females: 46 |
| Boone, 1992^52^ | C | USA | Healthy participants | 100 | 62.8 | 36 | NA | NA | NA | 64 | NA | NA | NA |
| Tupler, 1992^53^ | C | USA | Controls  DWMH | 66 | 61.8 | 15  9 | NA | NA | NA | 33  9 | NA | NA | NA |
| Rao, 1989^54^ | C | USA | Controls  LA | 50 | 43.7 | 10  1 | NA | NA | NA | 30  9 | NA | NA | NA |
| Jin, 2020^55^ | H | China | SIVD | 73 | 48.0 | 48 | NA | NA | NA | 25 | NA | NA | NA |
| Zhou, 2020^56^ | H | China | LI grade 0  LI grade 1  LI grade 2  LI grade 3 | 175 | 76.36 | 22  24  26  22 | NA | NA | NA | 23  20  21  17 | NA | NA | NA |
| Jokumsen-Cabral, 2019^57^ | H | Portugal | Controls  CADASIL | 47 | 57.85 | 6  11 | NA | NA | NA | 14  16 | NA | NA | NA |
| Kate, 2019^58^ | H | Canada | ICH | 71 | 69 | 52 | NA | NA | NA | 19 | NA | NA | NA |
| Liang, 2019a^59^ | H | Hong Kong | First-ever IS with lacunae  First-ever IS with CMBs | 195 | 66.17 | 73  54 | 64.9 | 59.5 | 59.1 | 44  24 | 68.0 | 72.6 | 7.5 |
| Liang, 2019b^60^ | H | China | Controls  LS | 831 | NA | 238  245 | NA | NA | NA | 187  161 | NA | NA | NA |
| Ling, 2019^61^ | H | France and Germany | CADASIL | 160 | 41.0 | 76 | NA | NA | NA | 84 | NA | NA | NA |
| Liu, 2019a^62^ | H | China | Controls  SIVD without CI  SIVD with CI | 81 | 69.54 | 10  16  16 | NA | NA | NA | 17  9  13 | NA | NA | NA |
| Liu 2019b^63^ | H | China | Subcortical infarcts | 50 | 52.6 | 30 | NA | NA | NA | 20 | NA | NA | NA |
| Liu, 2019c^64^ | H | China | Controls  SVD without CI  SVD with CI | 66 | 64.36 | 13  10  10 | NA | NA | NA | 12  11  10 | NA | NA | NA |
| Manso-Calderón, 2019^65^ | H | Spain | SVaD | 184 | 80.3 | 82 | NA | NA | NA | 102 | NA | NA | NA |
| Oudeman, 2019^66^ | H | Netherlands | VaCI | 58 | 68.9 | 35 | NA | NA | NA | 23 | NA | NA | NA |
| Reginold, 2019^67^ | H | Canada | WMH | 31 | 70.5 | 17 | NA | NA | NA | 14 | NA | NA | NA |
| Rudilosso, 2019^68^ | H | Spain | Subcortical infarcts | 67 | 66.3 | 40 | NA | NA | NA | 27 | NA | NA | NA |
| Staszewski, 2019^69^ | H | Poland | LS  VaD | 102 | 63.88 | 35  13 | NA | NA | NA | 17  37 | NA | NA | NA |
| Tsai, 2019^70^ | H | Taiwan | ICH | 257 | 63.15 | 162 | NA | NA | NA | 95 | NA | NA | NA |
| Wu, 2019^71^ | H | China | SIVD | 73 | 65.71 | 48 | NA | NA | NA | 25 | NA | NA | NA |
| Yu, 2019^72^ | H | Canada | Controls  SIVD | 54 | 71.7 | 11  14 | NA | NA | NA | 14  15 | NA | NA | NA |
| Zhang, 2019^73^ | H | China | Controls  Amnesic MCI with high grade WMH | 186 | 67.61 | 46  44 | NA | NA | NA | 44  52 | NA | NA | NA |
| Ishibashi, 2018^74^ | H | Japan | MCI without WMH  MCI with WMH | 75 | 78.1 | 11  25 | NA | NA | NA | 18  21 | NA | NA | NA |
| Kim, 2018^75^ | H | USA | Controls  Subcortical VaCI | 80 | 77.73 | 9  17 | NA | NA | NA | 10  44 | NA | NA | NA |
| Lisiecka-Ford, 2018^76^ | H | UK | SVD corresponding LS and Faz ≥ 2 | 114 | 70 | 75 | NA | NA | NA | 39 | NA | NA | NA |
| Anor, 2017^77^ | H | Canada | VaD | 34 | 75.3 | 16 | NA | NA | NA | 18 | NA | NA | NA |
| Yuan, 2017^78^ | H | China | Controls  LA | 100 | 70.45 | 19  21 | NA | NA | NA | 31  29 | NA | NA | NA |
| Zhong, 2017^79^ | H | China | WMH | 75 | 67.05 | 36 | NA | NA | NA | 39 | NA | NA | NA |
| Bella, 2016^80^ | H | Italy | Controls  VaCI | 45 | 66.21 | 9  10 | 66.05 | NA | NA | 11  15 | 66.27 | NA | NA |
| Hashimoto, 2016^81^ | H | Japan | CMBs median < 5  CMBs median > 5 | 22 | 69 | 5  8 | NA | NA | NA | 5  4 | NA | NA | NA |
| Hsu, 2016^82^ | H | Taiwan | Controls  VaMCI | 50 | 66.32 | 10  14 | NA | NA | NA | 20  6 | NA | NA | NA |
| Turk, 2016^83^ | H | Slovenia | Controls  Ischemic LA | 93 | 53.55 | 22  29 | NA | NA | NA | 18  24 | NA | NA | NA |
| Brookes, 2015^84^ | H | UK | Controls  Lacunar syndrome | 499 | 62.89 | 164  133 | NA | NA | NA | 139  63 | NA | NA | NA |
| Hsu, 2015^85^ | H | Taiwan | MCI | 31 | 75.4 | 19 | NA | NA | NA | 12 | NA | NA | NA |
| Brookes, 2014^86^ | H | UK | Controls  Lacunar syndrome plus LI | 125 | 68.68 | 36  25 | NA | NA | NA | 44  20 | NA | NA | NA |
| Delrieu, 2014^87^ | H | Multicentre | MCI | 65 | 74.8 | 48 | NA | NA | NA | 17 | NA | NA | NA |
| Ledesma-Amaya, 2014^88^ | H | Mexico | Controls  LI | 32 | 63.88 | 10  10 | NA | NA | NA | 6  6 | NA | NA | NA |
| Pinkhardt, 2014^89^ | H | Germany | DWMH Faz 1 ^a^  DWMH Faz 2  DWMH Faz 3 | 25 | 74.68 | 2  3  3 | 73.88 | NA | NA | 2  9  6 | 75.06 | NA | NA |
| Zi, 2014^90^ | H | China | Controls  PWMH | 32 | 61.73 | 7  7 | NA | NA | NA | 9  9 | NA | NA | NA |
| Deguchi, 2013^91^ | H | Japan | Controls  LI | 181 | 72.18 | 60  50 | NA | NA | NA | 45  26 | NA | NA | NA |
| Fang, 2013^92^ | H | China | Controls  SI  CMB  Both SI and CMB | 227 | 71.30 | 47  20  24  26 | NA | NA | NA | 44  26  17  23 | NA | NA | NA |
| Kim, 2013^93^ | H | USA | Subcortical VaMCI  Subcortical VaD | 127 | 73.8 | 23  28 | NA | NA | NA | 36  40 | NA | NA | NA |
| Narasimhalu, 2013^94^ | H | Singapore | LS without subjective CI  LS with subjective CI | 97 | 53 | 21  50 | NA | NA | NA | 9  17 | NA | NA | NA |
| Sudo, 2013^95^ | H | Brazil | Controls  VaMCI | 26 | 73.11 | 3  6 | NA | NA | NA | 8  9 | NA | NA | NA |
| Van Norden, 2013^96^ | H | Netherlands | SVD without subjective CI  SVD with subjective CI | 497 | 65.6 | 30  251 | NA | NA | NA | 17  199 | NA | NA | NA |
| Li, 2012^97^ | H | China | Controls  Ischemic LA | 40 | 65.45 | 12  11 | NA | NA | NA | 8  9 | NA | NA | NA |
| Quinque, 2012^98^ | H | Germany | Controls  Early cerebral microangiopathy | 32 | 64.42 | 13  7 | NA | NA | NA | 8  4 | NA | NA | NA |
| Yi, 2012^99^ | H | China | Controls  Subcortical VaMCI | 54 | 67.21 | 12  11 | NA | NA | NA | 16  15 | NA | NA | NA |
| Fernández, 2011^100^ | H | Spain | Controls  Subcortical VaMCI | 38 | 71.5 | 9  13 | NA | NA | NA | 10  6 | NA | NA | NA |
| Xiong, 2011^101^ | H | China | LS without cognitive complaints  LS with cognitive complaints | 75 | 70.75 | 20  19 | NA | NA | NA | 23  13 | NA | NA | NA |
| Hassan, 2010^102^ | H | Egypt | Controls  LI | 42 | 58.59 | 8  18 | NA | NA | NA | 4  12 | NA | NA | NA |
| Pascual, 2010^103^ | H | Spain | Controls  VWMD without dementia  VWMD with dementia | 36 | 80.1 | 6  6  6 | NA | NA | NA | 6  6  6 | NA | NA | NA |
| Seo, 2010^104^ | H | South Korea | Controls  Subcortical VaMCI  Subcortical VaD | 150 | 69.22 | 42  19  9 | NA | NA | NA | 54  15  11 | NA | NA | NA |
| Staekenborg, 2010^105^ | H | Multicentre | VaD | 401 | 73 | 247 | NA | NA | NA | 154 | NA | NA | NA |
| Price, 2009^106^ | H | USA | Controls  Dementia with mild LA  Dementia with moderate LA  Dementia with severe LA | 168 | 78.89 | 8  13  15  5 | NA | NA | NA | 16  60  29  22 | NA | NA | NA |
| Zhou, 2009^107^ | H | China | Controls  MCI from SVD | 136 | 67.06 | 45  36 | NA | NA | NA | 35  20 | NA | NA | NA |
| Gainotti, 2008^108^ | H | Italy | Controls  MCI with multiple SI | 108 | 71.24 | 37  26 | NA | NA | NA | 28  17 | NA | NA | NA |
| Nordlund, 2007^109^ | H | Sweden | Controls  VaMCI | 120 | 66.75 | 28  22 | NA | NA | NA | 32  38 | NA | NA | NA |
| Nordahl, 2005^110^ | H | USA | Controls  MCI with severe WMH | 28 | 78.25 | 3  5 | NA | NA | NA | 14  6 | NA | NA | NA |
| Van Zandvoort, 2005^111^ | H | Netherlands | Controls  LI | 38 | 62.06 | 8  16 | NA | NA | NA | 4  10 | NA | NA | NA |
| Garrett, 2004^112^ | H | USA | Controls  VaCI  VaD | 69 | 77.22 | 11  10  17 | NA | NA | NA | 14  8  9 | NA | NA | NA |
| Graham, 2004^113^ | H | UK | Controls  Subcortical VaD | 38 | 69.65 | 9  14 | NA | NA | NA | 10  5 | NA | NA | NA |
| Van Zandvoort, 2003^114^ | H | Netherlands | LI ^a^ | 17 | 60.3 | 12 | 61.17 | NA | NA | 5 | 58.2 | NA | NA |
| Kramer, 2002^115^ | H | USA | Controls  SIVD | 39 | 73.08 | 12  6 | NA | NA | NA | 15  6 | NA | NA | NA |
| Maeshima, 2002^116^ | H | Japan | Controls  Periventricular WMH | 84 | 48.82 | 32  6 | NA | NA | NA | 38  8 | NA | NA | NA |
| Yuspeh, 2002^117^ | H | USA | Controls  SIVD | 67 | 73.82 | 24  19 | NA | NA | NA | 14  10 | NA | NA | NA |
| Aharon-Peretz, 2000^118^ | H | Israel | VaD | 30 | 71.75 | 20 | NA | NA | NA | 10 | NA | NA | NA |
| Yamauchi, 2000^119^ | H | Japan | Controls  LI | 62 | 68.37 | 13  19 | NA | NA | NA | 21  9 | NA | NA | NA |
| Binetti, 1995^120^ | H | Italy | MID | 32 | 76.1 | 17 | NA | NA | NA | 15 | NA | NA | NA |
| Lewine, 1993^121^ | H | USA | Controls  WMH | 20 | 35.2 | 4  4 | 34.85 | NA | NA | 6  6 | 43.3 | NA | NA |
| Johansson, 2020^122^ | M | Netherlands | Controls  MCI | 157 | 71.0 | 52  31 | NA | NA | NA | 52  22 | NA | NA | NA |
| Atwi, 2018^123^ | M | Canada | Controls  WMH Faz > 2 | 37 | 47.86 | 9  8 | NA | NA | NA | 10  10 | NA | NA | NA |
| Gonçalves, 2017^124^ | M | Portugal | Controls  SVaD | 56 | 75.9 | 19  10 | NA | NA | NA | 21  6 | NA | NA | NA |

Healthy participants included those defined as neurologically, functionally or cognitively healthy or community-dwelling individuals. Total participants refer to the relevant populations extracted for this work. ^a^ Studies that provided more stratified information of SVD groups than controls, so controls were not included in this review. Abbreviations: AIS = acute ischemic stroke, CADASIL = cerebral autosomal dominant arteriopathy with subcortical infarcts and leukoencephalopathy, CMB = cerebral microbleeds, CI = cognitive impairment, DWMH = deep white matter hyperintensities, Faz = Fazecas score, ICH = intracerebral hemorrhage, IS = ischemic stroke, LA = leukoaraiosis, LI = lacunar infarct, LS = lacunar stroke, MCI = mild cognitive impairment, MID = multi-infarct dementia, NA = not available, SI = silent brain infarcts, SIVD = subcortical ischemic vascular dementia, SVD = cerebral small vessel disease, VaCI = vascular cognitive impairment, VaD = vascular dementia, VaMCI = vascular mild cognitive impairment, VWMD = vascular white matter disease, WMH = white matter hyperintensities, WML = white matter lesions.

**Supplementary Table S2. Results of quantitative tests excluding studies with quality scores < 5.5/8.**

| **Analysis** | **Statistical test** | **Results** |
| --- | --- | --- |
| Trends across study settings: comparisons of sex ratio per study type. | Kruskal-Wallis test, pairwise Mann-Whitney U-tests and Bonferroni post-hoc correction. | Significant differences were in sex ratios across study setting (H = 17.31, df = 2, p < 0.001). Greater sex ratio in HB compared with CB studies (p_corrected_ < .001). |
| Correlation between the deviation of the sex ratio and the size of the recruited population. | Spearman’s rank correlation coefficient. | Negative correlation in CB (rho_Spearman_ = -0.44, p = 0.006) and no correlation in HB studies (rho_Spearman_ = .028, p = 0.87). |
| Trends across time: comparisons between recent and previously published research. | Mann-Whitney U-test. | No significant differences between sex ratios of recent studies compared with those previously published (U = 715, p = 0.80). This finding was consistent after classifying by study type (U = 230, p = 0.11 in CB; U = 143, p = 0.42 in HB). |
| Trends across SVD severity: Comparisons between healthy to mild non-clinical SVD and moderate to severe SVD. | Mann-Whitney U-test. | Sex ratio in moderate to severe SVD was greater than in healthy to mild SVD (U = 1043.5, p < 0.001). |
| Trends across SVD presentation: Comparisons between healthy to mild non-clinical SVD, moderate to severe non-clinical SVD, cognitive SVD and cerebrovascular SVD. | Kruskal-Wallis test, pairwise Mann-Whitney U-tests and Bonferroni post-hoc correction. | Significant differences in sex ratios across SVD presentation (H = 28.06, df = 3, p < 0.001). Participants with stroke presentations showed a greater sex ratio, when compared with healthy to mild SVD (p_corrected_ < .001), cognitive SVD (p_corrected_ = .002), and moderate to severe non-clinical SVD (p_corrected_ = .001). |

Abbreviations: CB = community-based, HB = hospital-based, SVD = cerebral small vessel disease.
